# Occupational and environmental exposure to SARS-CoV-2 in and around infected mink farms

**DOI:** 10.1101/2021.01.06.20248760

**Authors:** Myrna M.T. de Rooij, Renate W. Hakze-Van der Honing, Marcel M. Hulst, Frank Harders, Marc Engelsma, Wouter van de Hoef, Kees Meliefste, Sigrid Nieuwenweg, Bas B. Oude Munnink, Isabella van Schothorst, Reina S. Sikkema, Arco N. van der Spek, Marcel Spierenburg, Jack Spithoven, Ruth Bouwstra, Robert-Jan Molenaar, Marion Koopmans, Arjan Stegeman, Wim H.M. van der Poel, Lidwien A.M. Smit

## Abstract

Unprecedented SARS-CoV-2 infections in farmed minks raised immediate concerns regarding human health which initiated intensive environmental investigations. Air sampling was performed in infected mink farms, at farm premises and at residential sites. A range of other environmental samples were collected from minks’ housing units including bedding material. Inside the farms, high levels of SARS-CoV-2 RNA were found in airborne dust, on surfaces, and on various other environmental matrices. This warns for occupational exposure which was substantiated by considerable SARS-CoV-2 RNA concentrations in personal air samples. Dispersion of SARS-CoV-2 to outdoor air was found to be limited and SARS-CoV-2 RNA was not detected in air samples collected beyond farm premises, implying a negligible environmental exposure risk for nearby communities. Our occupational and environmental risk assessment is in line with whole genome sequences analyses showing mink-to-human transmission in farm workers, but no indications for direct zoonotic transmission events to nearby communities.

## Introduction

The COVID-19 pandemic has its grip on the world. After the initial jump from an animal reservoir or as yet unknown intermediate animal host to humans, human-to-human transmission took off. Several incidental cases have been reported of transmission from humans to animals^1^ but large outbreaks amongst commercially kept animals were not reported until minks (*Neovison vison*) at fur farms were found to be infected in April 2020^2^. Initially SARS-CoV-2 infections in mink farms were only observed in the Netherlands, but over time also in the U.S., Denmark and other European countries^3–5^. SARS-CoV-2 outbreaks in mink farms and transmission experiments with ferrets ^6–8^ show that SARS-CoV-2 is efficiently transmitted between minks and related species.

Occurrence of unprecedented SARS-CoV-2 infection in farmed minks raised immediate concerns regarding human health, both for the farm worker population, for the general population living in the vicinity of the infected farms and in view of global health considering the potential evolution of SARS-CoV-2 by large scale animal passaging. Therefore, a One-Health approach was deployed to gain insight in the (genomic) epidemiology, sources and modes of transmission of the outbreaks in the mink farms, and the associated human health risks^2,9,10^. Using whole genome sequencing, evidence was provided of transmission within farms of SARS-CoV-2 from humans to minks, between minks, and from minks back to humans^9^. Both in The Netherlands^9^ and Denmark^11^, widespread transmission between farms occurred, with yet unknown mode of transmission.

In this paper, we focus on intensive environmental investigations performed in the first SARS-CoV-2 infected mink farms notified in The Netherlands, and in their surroundings to better understand occupational and environmental health risks for workers and neighbouring residents. Study objectives were to:

1. Measure levels of SARS-CoV-2 in air in mink farms including personal exposure using portable samplers
2. Study potential dispersion of SARS-CoV-2 to the ambient environment by means of outdoor air sampling
3. Assess SARS-CoV-2 contamination of surfaces and materials sampled from the minks’ housing units

## Methods

### Investigated farms

On April 23^rd^, SARS-CoV-2 infection was established in the first Dutch farm, NB1, which has two separate locations, NB1A and NB1B, which are 115m apart. Two days later, animals from another farm tested positive, NB2, situated at 14 km distance from NB1. Both NB1 and NB2 experienced increased mortality amongst the minks coinciding with respiratory signs starting in the first half of April. Serological findings and sequencing of viral genomes suggested that virus circulation had been ongoing for some time before detection^2,9,10^. From April 28 onwards, environmental sampling started at and around NB1 and NB2. On May 6^th^, SARS-CoV-2 infection was detected in animals from two additional farms, NB3 and NB4. NB4, in contrast to the other farms, appeared to be more recently infected at the time of detection, based on disease history, serology and sequence diversity^2,9,10^. Therefore, NB4 was included for environmental sampling from May 13^th^ onwards. See Supplemental Methods A for additional information on the outbreaks including a timeline and background information on husbandry practices in general.

### Design of environmental sampling

We conducted long-term air sampling to detect potential dispersion of SARS-CoV-2 to the outdoor environment and potential exposure at nearby residential areas. As levels in the outdoor environment were not expected to be high, this involved continuous multi-day sampling in order to measure high volumes of air per sample. Long-term air samples were collected at both farms’ premises and at residential sites.

In addition to long-term sampling, we focused on sampling inside the farm, including sampling of air, settling dust, and surfaces and materials sampled from minks’ housing units. Upwind and downwind sampling around the mink houses was performed to assess dispersion. For this part of the study, we visited each farm once per week during three weeks (T1, T2, T3). In a later phase of the study, when the more recent outbreak at NB4 became apparent, long-term outdoor air sampling at NB4 was performed during three weeks, and sampling inside the farm was done once. On June 3^rd^, the Dutch government decided to cull all minks on all infected farms. To gain insight in potentially remaining environmental contamination after culling, NB4 was revisited 14 days post-culling, to collect environmental samples of minks’ housing units.

### Air sampling: PM_10_ and inhalable dust

Air sampling was performed by means of filter-based techniques using teflon filters (Pall Corporation, Ann Arbor, USA). With this technique, air is forced through a filter and particles present in the air, including potentially SARS-CoV-2 contaminated particles, are captured. To gain insight in potential differences related to particle size fraction, all air sampling was performed in parallel to measure both Particulate Matter 10 (PM_10_) and inhalable dust based on the by European norm (EN) defined size fractions^12,13^. PM_10_ is defined by particles of a nominal aerodynamic diameter of 10 μm or less, particles of this size or smaller can penetrate the tracheobronchial regions of the airways.

Inhalable dust includes PM_10_ but also larger particles (typically up to 100 μm) and is defined by particles small enough to enter the respiratory tract.

### Multiple-day outdoor air sampling

At the infected farms, consecutive 3-to 5-day sampling of outdoor air was performed at a central position within 10m from the (open) wall of the farm. Both sampling of PM_10_ and inhalable dust was performed in parallel at 1.50m height (average breathing height of humans), for technical details see Supplemental Methods B. After the first measurement week at NB4, measurement equipment was installed at three additional locations (B,C,D) within short distances from the initial location (A), where positive samples had been detected, see Supplemental Figure S1 for a map of the measurement locations. At B, C and D, total suspended particles (larger size fraction than inhalable dust) were additionally sampled.

Three residential sites were included in 7-day air sampling. Three consecutive measurements per site were performed in the vicinity (1500 m) of farms NB1A and NB1B and in the vicinity (1200 m) of farm NB2. In addition, measurements were also performed at a residential site in a mink-free area (>70 km from NB1 and NB2) as a background location.

### Sampling inside farm and downwind/upwind

#### Air sampling

Six-hour stationary air sampling and 8-hour personal air sampling were performed for both PM_10_ and inhalable dust in parallel, for technical details and pictures see Supplemental Methods B. Personal air samples were collected using portable pumps and attaching the sampling heads within the breathing zone of the fieldworker. Stationary air sampling was performed by attaching sampling heads side-by-side at 1.50m height. Stationary air sampling was performed at three spots distributed within the farm which remained the same over time. The locations of the six-hour air sampling outside the farm were based on the wind direction of the measurement day. Upwind sampling was performed at 50 meter distance from the farm, and downwind sampling at 10-20 meter and 100 meter distance.

#### Settling dust sampling

Settling dust sampling was performed by using Electrostatic Dust fall Collectors (EDCs), which are electrostatic cloths placed in a disposable holder^14^. EDCs were placed at 11 spots distributed throughout the farm. EDCs were placed in proximity (<0.40m) of the minks by placing them on the unused top layer of the mink’s housing units (1.60m height). At farm NB1A and NB1B, also some EDCs could be placed further away from minks (several meters) by placing these onto hanging plates or on a stand positioned in an empty alley (see Supplemental Methods B for pictures). After one week, the exposed EDCs were collected and replaced by new EDCs.

#### Sampling of minks’ housing units

Per farm visit, a minimum of 10 housing units were sampled, including those of recently deceased minks (<2 days), and at least 3 alive minks (see Supplemental Methods B). Swipes were collected of the material settled on the hardboard border on the front of the housing unit. Bedding material, consisting of straw/hay, was collected from the night/nest box. Food residues were scraped off the top of the cage where minimally once a day fresh food is placed. Swabs were taken of the rim of the drinker cup. Faecal material was collected from the inside of the cage when present, or from the floor beneath the cage.

### Procedures and laboratory analyses

All fieldworkers wore PPE including full face mask and remained SARS-CoV-2 negative. No positive field blanks were identified throughout the study. Each fieldwork day, all samples were immediately stored after collection at 4C° and directly brought to a BSL-2 lab where samples were prepared for storage at -80°C the same day. Samples were transported in batches on dry ice to another BSL-2 lab for RNA extraction and qPCR analyses on SARS-CoV-2. See Supplemental Methods C for details about laboratory procedures.

### Data processing and statistics

To enable quantitative analyses, viral load in the positive samples was computed and expressed in number of copies per standard unit (e.g. per cubic meter of air; see Supplemental Methods C).

Multivariable linear regression or logistic regression (viral presence versus absence) analyses were performed to explore associations with determinants. For all models, assumptions were checked including distribution of residuals. To meet assumptions for linear modelling, viral load was log-transformed because of the otherwise skewed distribution and samples testing negative were assigned an arbitrary value (1/10^th^ of the limit of detection).

## Results

### Air samples

#### Farms NB1A, NB1B, NB2

At the first farm visit, SARS-CoV-2 RNA was detected inside each farm in one out of three 6-hour inhalable dust samples (see Table 1). At farm NB1B during the first visit also personal sampling was performed of which one of the two 8-hour inhalable dust samples was positive for SARS-CoV-2 RNA. Quantification of these four positive active air samples showed concentrations of around 4 x 10^3^ RNA copies/m^3^ (Ct values 35 to 36). All of the parallel collected PM_10_ samples inside the farm and all other inhalable dust and PM_10_ air samples inside or near the mink houses were negative.

**Table 1.**
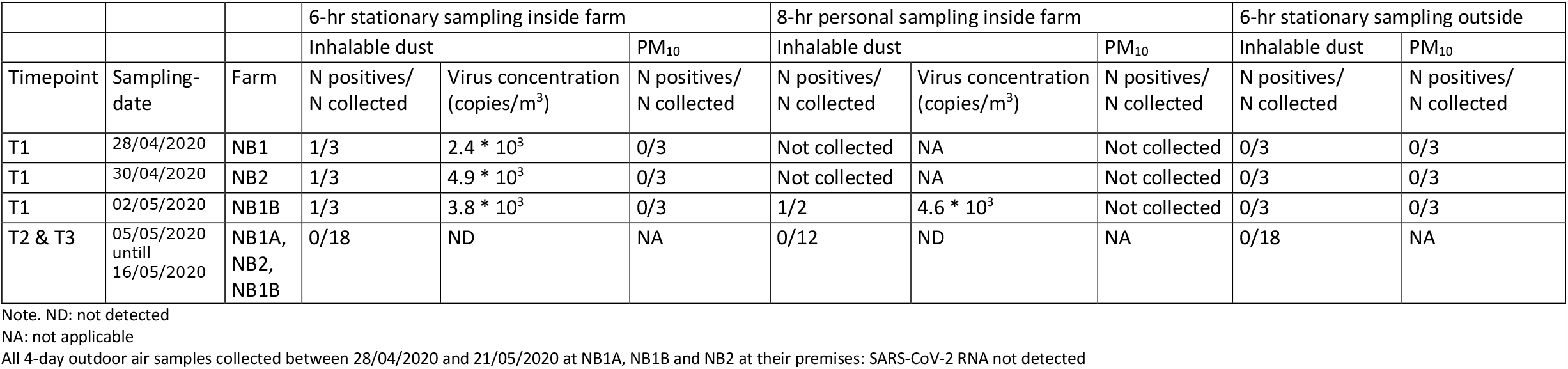
Overview of SARS-CoV-2 RNA in PM_10_ and inhalable dust samples collected at mink farms in a later phase of the ongoing SARS-CoV-2 outbreak

SARS-CoV-2 RNA was detected in a high number of settled dust samples (75 out of 92, 82%). All EDCs deployed at the first farm visit were positive (range in Ct values 25.1 to 34.6) at all three farms with viral RNA loads ranging from 4 x 10^3^ to 7 x 10^3^ copies/m^2^ per day sampled (see Figure 1). For NB2 all EDCs deployed at the second and third farm visit were also all positive, for T2 (one week after T1) at NB1A and NB1B the percentage positives dropped to 73% and 80%, respectively and further to 64% and 27% for T3. Results of the multivariable modelling (see Supplemental Table S1) showed a significant (p<0.05) decrease in viral RNA load over time at all three farms (factor 4 to 5 difference per week). Furthermore viral RNA loads in EDCs placed in very close proximity to minks were on average 3 times higher than those further away from animal cages (p<0.05).

**Figure 1.**
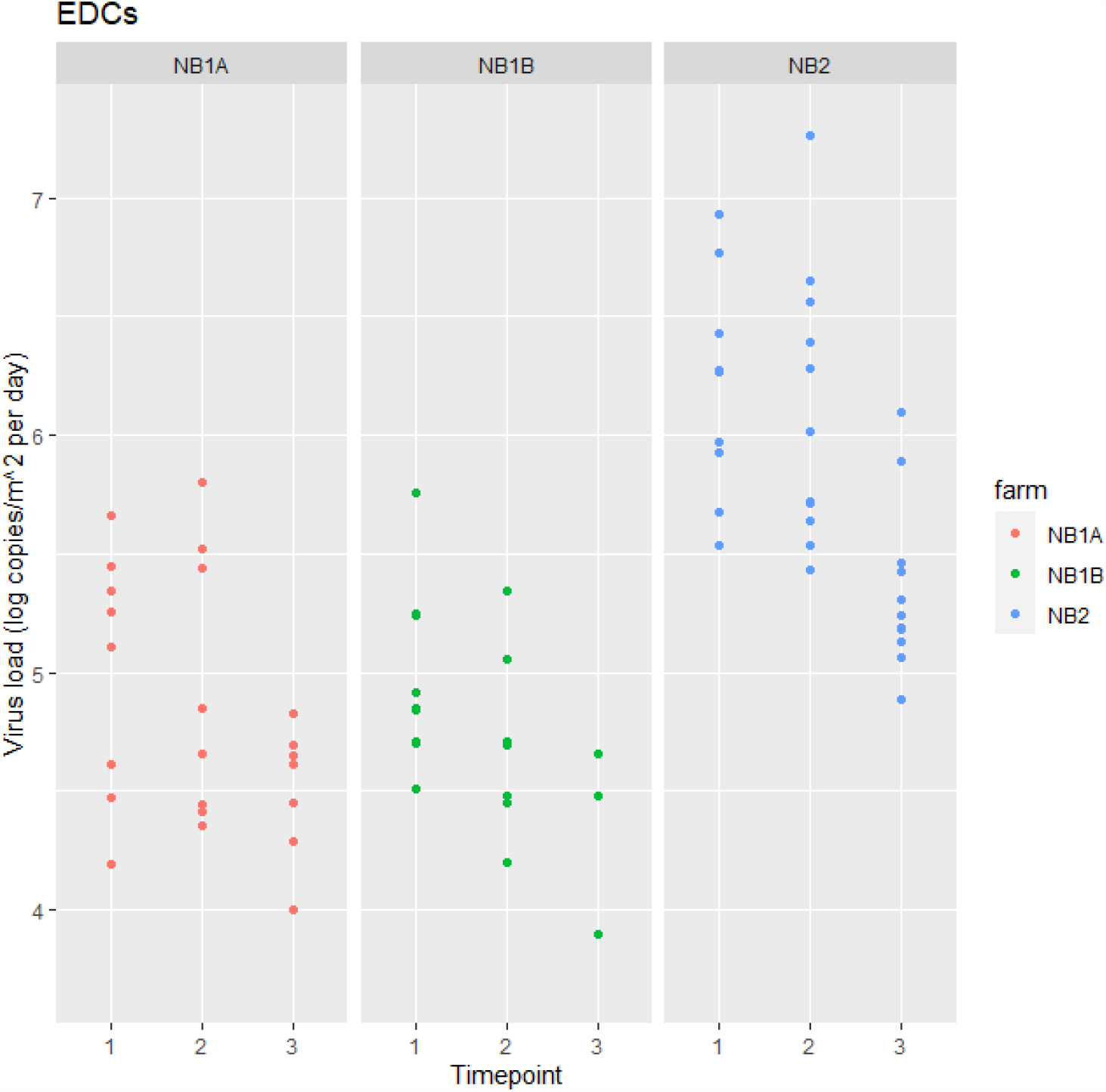
Overview of viral load in settling dust samples per farm over time (detectable levels are plotted, percentages of non-detects are noted below) Note. NB1A – T2: 3/11 (27%) < LOD ; T3: 4/11 (36%) < LOD NB1B – T2: 2/10 (20%) < LOD; T3: 8/11 (73%) < LOD NB2 – none < LOD

#### Residential sites

All of the consecutively collected 7-day ambient air samples of PM_10_ at the three residential sites (three samples per site) were negative for SARS-CoV-2.

#### Farm NB4

Of the air samples collected in farm NB4, in a more early phase of the outbreak than NB1 and NB2, three out of six 6-hour inhalable dust samples and both 8-hour personal inhalable dust samples were positive, see Table 2. Concentrations measured in the personal air samples were higher compared to the stationary air samples, roughly 4 x 10^5^ copies/m^3^ (Ct 31.7/31.8) compared to 7 x 10^3^ to 2 x 10^4^ (range in Ct values 33.0 to 34.4). Two out of six stationary PM_10_ samples and both personal PM_10_ samples were positive, with lower concentrations compared to the inhalable dust samples.

**Table 2.** Overview of SARS-CoV-2 RNA in PM_10_ and inhalable dust samples collected at a mink farm (NB4) in a more acute phase of SARS-CoV-2 outbreak

| Sampling-date | Measurement | Inhalable |  | PM<sub>10</sub> |  | Total suspended particles |  |
| --- | --- | --- | --- | --- | --- | --- | --- |
|  |  | Negative sample(s) | Positive sample(s) - virus concentration (copies/m<sup>3</sup>) | Negative sample(s) | Positive sample(s) - virus concentration (copies/m<sup>3</sup>) | Negative sample(s) | Positive sample(s) - virus concentration (copies/m<sup>3</sup>) |
| 13/05/2020 | Multiple-day outdoor: spot A only | none | A: 7.1 * 10<sup>2</sup> | A | none | Not collected | Not collected |
| 16/05/2020 | Multiple-day outdoor: spot A only | none | A: 3.6 * 10<sup>3</sup> | A | none | Not collected | Not collected |
| 19/05/2020 | 6-hr indoor stationary (sampling spots I, II, III, IV, V, VI) | I; II; V | III: 6.6 * 10<sup>3</sup><br>IV: 9.8 * 10<sup>3</sup><br>VI: 1.9 * 10<sup>4</sup> | I; II; III; V | IV: 5.5 * 10<sup>3</sup><br>VI: 2.5 * 10<sup>3</sup> | Not collected | Not collected |
|  | 8-hr indoor personal (person X and person Y) | none | X: 4.5 * 10<sup>4</sup><br>Y: 4.3 * 10<sup>4</sup> | None | X: 2.0 * 10<sup>4</sup><br>Y: 3.0 * 10<sup>3</sup> | Not collected | Not collected |
| 21/05/2020 | Multiple-day outdoor: expanded (spot A + B,C,D) | none | A: 2.6 * 10<sup>3</sup> | none | A: 1.1 * 10<sup>2</sup> | A; C; D | B: 6.9 * 10<sup>2</sup> |
| 25/05/2020 | Multiple-day outdoor: expanded (spot A + B,C,D) | A; B; C; D | none | A | none | B; D | A: 4.5 * 10<sup>2</sup><br>C: 3.1 * 10<sup>2</sup> |
| 28/05/2020 | Multiple-day outdoor: expanded (spot A + B,C,D) | A; D | none | none | A: 1.2 * 10<sup>3</sup> | D | A: 4.8 * 10<sup>3</sup><br>B: 1.7 * 10<sup>3</sup><br>C: 2.7 * 10<sup>2</sup> |

Note. I, II, III, IV, V, VI: stationary indoor air sampling spots, see Supplemental Figure S1 for a map of the lay-out of the farm and measurement spots
A, B, C, D: stationary outdoor air sampling spots, see Supplemental Figure S1 for a map of the lay-out of the farm and measurement spots
X: personal air sampling of fieldworker X
Y: personal air sampling of fieldworker Y

In 4-day outdoor air samples collected at the premises, SARS-CoV-2 RNA was detected at measurement location A positioned within 1.5-meters of the farm’s open entrance, but also at measurement spots B and C positioned tens of meters from the farm’s open entrance. PM_10_ samples collected at spot A were either negative or contained limited levels of SARS-CoV-2 RNA. Inhalable dust and total suspended particles samples contained levels of SARS-CoV-2 RNA, ranging from 3 x 10^2^ to 5 x 10^3^ copies/m^3^ (range in Ct values 32.3 to 35.0). At location D, 20 meters from the entrance, all samples were negative.

### Samples of minks’ housing units

#### Pre-culling

SARS-CoV-2 RNA was detected in all swipes of the minks’ housing units (N=99, lowest Ct value 21.5) collected pre-culling. Also a high percentage of the bedding material samples (83%, 78 out of 94, lowest Ct value 15.9) was positive and to a lesser extent the samples of faecal material (54%, 51 out of 95, lowest Ct value 24.6). Some of the swabs of the drinker cups and few samples of the food residue were positive (31%, 30 out of 97, lowest Ct value 24.8; 10%, 9 out of 90, lowest Ct value 28.9; respectively). Significant differences in viral load in the swipes and bedding material samples were observed between farms, see Table 3 and Supplemental Figure S2. Viral load was estimated to be 7 times higher in swipes and 50 times higher in bedding material samples for NB4 compared to NB1A and even larger differences when compared to NB1B and NB2. Higher viral loads were observed in bedding material of housing units belonging to minks that were recently deceased versus minks still alive. Significantly lower odds of detection in faecal material and swabs of drinker cups was observed for sampling later in time and differences between farms were observed with significant lower odds of detection at NB2 (Table 3). For swabs of the drinker cups, higher odds of detection was also associated with recent decease of the mink.

**Table 3.** Results of multivariable modelling on SARS-CoV-2 RNA in the different sample types of mink housing units

|  | Virus load (log copies) |  |  |  |  |  | Virus presence (non-detect/detect) |  |  |  |  |  |
| --- | --- | --- | --- | --- | --- | --- | --- | --- | --- | --- | --- | --- |
|  | Swipe |  |  | Bedding material |  |  | Faecal material |  |  | Swab drinker cup |  |  |
| Variable | Ratio | 95% CI LB | 95% CI UB | Ratio | 95% CI LB | 95% CI UB | OR | 95% CI LB | 95% CI UB | OR | 95% CI LB | 95% CI UB |
| Farm NB1A (indicator) | . | . | . | . | . | . | . | . | . | . | . | . |
| Farm NB1B | 0.43* | 0.22 | 0.83 | 2.00 | 0.24 | 16.0 | 0.31 | 0.081 | 1.11 | 0.28 | 0.061 | 1.09 |
| Farm NB2 | 0.37* | 0.20 | 0.70 | 0.18 | 0.02 | 1.50 | 0.16* | 0.042 | 0.56 | 0.15* | 0.031 | 0.58 |
| Farm NB4 | 6.80* | 2.70 | 17.0 | 51.0* | 2.60 | 1000 | 0.19 | 0.025 | 1.28 | 1.92 | 0.296 | 14.32 |
| Timepoint 1 (indicator) | . | . | . | . | . | . | . | . | . | . | . | . |
| Timepoint 2 | 1.10 | 0.58 | 2.10 | 2.10 | 0.22 | 15.0 | 0.4 | 0.115 | 1.28 | 0.25* | 0.058 | 0.97 |
| Timepoint 3 | 0.57 | 0.30 | 1.10 | 1.10 | 0.12 | 8.80 | 0.11* | 0.027 | 0.38 | 0.22* | 0.045 | 0.88 |
| Mink recently deceased (versus live mink) | 1.60 | 0.92 | 2.60 | 58.0* | 10.0 | 340 | 0.47 | 0.161 | 1.31 | 7.76* | 2.156 | 38.32 |

Note. Number of samples (percentage above limit of detection) collected per type: swipe n= 99 (100%), bedding material n= 94 (83%), faecal material n= 95 (54%), swab drinker cup n= 97 (31%). Logistic regression was applied for faecal and drinker cup samples due to the lower number of quantifiable samples.
Ratio = estimate of association to the power 10 to represent ratio in viral load
OR = odds ratio
95% CI LB = lower bound of the 95% confidence interval
95% CI UB = upper bound of the 95% confidence interval
Marked \* then P-value < 0.05

#### Post-culling

Samples collected post-culling at NB4 of the same housing units sampled pre-culling showed a clearly decreased detection of viral RNA. However, RNA was still measurable in 14% of the swipes and 21% of faecal material, and especially in bedding material (top layer 57%, bottom layer 85%). A large drop in viral load detected in the top layer of the bedding material collected post-culling was observed compared to pre-culling (factor 100 difference, p-value<0.001), however the drop in viral load detected in the bottom layer was considerably less compared to pre-culling viral load (factor 10 difference, p-value<0.05) (see Supplemental Table S2 and Figure S3).

## Discussion

Air and surfaces in infected mink farms were found to be highly contaminated with SARS-CoV-2 RNA. Airborne inhalable dust contaminated with SARS-CoV-2 RNA was detected inside each investigated farm. The vast majority of the settling dust samples collected inside every farm contained SARS-CoV-2 RNA and also many of the samples of the minks’ housing units. In contrast to the high level of contamination detected inside the farm, in the outdoor air no SARS-CoV-2 RNA or incidental low concentrations were detected. This raises caution for health risks associated with working inside the farms, and suggests a negligible role for transmission of SARS-CoV-2 by dispersion via air to neighbourhoods of farms, respectively.

In the course of the outbreak, effective spread of SARS-CoV-2 amongst minks was observed and also transmission between minks and farm workers^2,9,10^. The high levels of environmental contamination inside the farm suggest a potential role of environmental exposure in mink-to-mink and mink-to-human transmission. The intensity of animal handling by farm workers varies over the course of a year and is typically high in April to June which is the period from the end of gestation until weaning. Given the high rate of animals tested positive^2,10^ and the viral loads detected in samples of their direct vicinity, animal handling is a likely cause of exposure for farm workers. Nevertheless, zoonotic transmission events were also observed in the months when no/little animal handling was performed^9^. SARS-CoV-2 contaminated airborne particles of sizes smaller than 10μm and also larger were detected in the air in the breathing zone of our fieldworkers (throughout the study confirmed negative SARS-CoV-2 status) who did not handle the animals but performed environmental sampling wearing PPE including full face mask. Both smaller and larger sized airborne particles should be considered relevant to health as research suggested that deposition of inhaled SARS-CoV-2 contaminated particles anywhere along the respiratory tract has the potential to initiate infection^15^. The RNA concentrations in inhalable dust samples were considerable. Only two personal air samples were eligible for viability testing based on Ct levels, but attempts made were unsuccessful. This was not surprising given the challenges with viability testing overall^16^, but also considering that the sampling strategy was not set up for this purpose. For air sampling in general, it is well-known that the sampling technique itself can have a considerable impact on pathogen viability making it challenging to actually assess the status of a pathogen when in the air^17,18^. Recently, research performed in experimental settings^19,20^ and in a hospital situation^21^ found viable SARS-CoV-2 in air samples. In aerosols generated under laboratory conditions, SARS-CoV-2 remained stable for several hours (<16 hours)^19,20^. These findings support the potential risk of infection due to transmission via air.

Airborne SARS-CoV-2 RNA could be a result of direct shedding into the air by an infectious mink (e.g. via sneezing, breathing) and/or indirectly via shedding into the environment and consequently becoming airborne of particles emanating from contaminated matrices (e.g. dust particles arising from bedding material). SARS-CoV-2 experiments performed with ferrets showed transmission via respiratory droplets, aerosols and/or fomites between ferrets placed apart at 10cm^6^ and >1meter^8^. In the infected mink farms, we performed stationary air sampling at a fixed 50cm distance from the nearest minks and personal air sampling naturally at varying distances up to as close as 10cm thus we could have detected direct shedding of respiratory droplets and/or aerosols.. The co-occurrence of the indirect route of airborne spread of contaminated particles is also highly likely, given the high levels of contamination observed in matrices easily becoming airborne like bedding material, and the presence of airborne dust generating events, such as uncontrolled air/wind flows, and movements of animals, food cars, and humans. Visual inspections of the settled dust collected by means of EDCs confirmed the high level of dust inside the mink farms. Occupational exposure inside the farm appeared to be highest during the acute phase of the outbreak amongst minks, as shown by SARS-CoV-2 RNA presence in settling dust even in empty rows thus at distances of several metres from the animals. Housing units of recently deceased minks were identified as hotspots for environmental contamination.

Environmental RNA load is the net result of shedding rates by SARS-CoV-2 infectious minks and processes like degradation, or removal/cleaning. Animal investigations showed a high seroprevalence amongst minks at the investigated farms^10^ indicating many of the minks had been infectious at some point in time and cleaning is performed only sporadically. The rate of RNA decay is influenced by factors like temperature and humidity, chemical exposure (e.g. reactive oxygen species, alkylating agents) and radiation exposure (e.g. ultraviolet radiation)^22^ and depending on the environmental matrix, more or less decay may take place. SARS-CoV-2 RNA was still detectable in the environment two weeks after all animals were culled, similar to what has been observed in an outbreak investigation study performed at a cruise ship^23^. The last time the minks’ housing units were cleaned was months before the start of the outbreak thus results were not affected by a cleaning regime. Proper cleaning/disinfecting of such a contaminated environment is difficult and warns for extended precaution in the period after culling. Actual characterization of the infection risk related to measured environmental contamination is hampered as no insight was gained in infectiousness of SARS-CoV-2 in the various matrices sampled. Experimental research is warranted to quantify the probability of infectiousness of SARS-CoV-2 over time while in the environment.

The unprecedented SARS-CoV-2 infections in farmed minks raised immediate concerns regarding public health which resulted in government imposed enclosure of the area around the farms awaiting results of outdoor air investigations. In contrast to the high environmental contamination detected indoor, little to no presence of SARS-CoV-2 RNA was detected just outside the mink houses. In none of the 54 outdoor air samples collected at NB1A, NB1B and NB2 was SARS-CoV-2 RNA detected. Considering the limit of detection (∼10 to 28 copies per cubic metre of air) implies that SARS-CoV-2 RNA was either not present in outdoor air, or if present then in very low concentrations. At NB4, the farm in a more acute phase of the SARS-CoV-2 outbreak, a positive outdoor air sample was detected which triggered expansion of the number of sampling spots. The outcomes clearly showed notable airborne RNA concentrations near the open entrance (<1.5m) and a considerable drop in concentrations several meters further. Outside the farm premises, at a distance of 20 meter from the minks, no RNA was detected. As observed indoors, also the outdoor air samples showed concentrations in PM_10_ to be clearly lower compared to inhalable dust/total suspended particles. Considering natural dispersion, large particles especially >30µm deposit quickly and typically do not reach distances further than tens of metres^24,25^. Particles sized 10µm or smaller, are able to disperse further but given the concentrations measured and the strong reduction over short distances observed, it is unlikely that a potential infection risk for the community exists at municipal roads near the farms. Especially when considering the outdoor situation, including various environmental factors unfavourable for viruses including UV-radiation. Consequently the direct environmental health risk for passers-by or neighbouring residents of the infected mink farms is expected to be negligible. This substantiates findings reported by Oude Munnink et al.^9^ which indicated no spill-over to people living in close proximity based on whole-genome sequencing of SARS-CoV-2 recovered from samples from minks and humans. Sequences of COVID-19-patients living in the vicinity of infected mink farms did not cluster with sequences identified in minks and mink farmers but reflected the general diversity seen in the national COVID-19-patient databases.

Outbreak investigations performed around infected mink farms in Denmark suggested that virus transmission to the local community was caused by social contacts of infected mink farmers and workers with others^11^. Our finding of ambient air posing a negligible risk of infection is not only relevant considering public health, but also suggests that transmission via air is an unlikely route for the widespread and still unexplained ongoing farm-to-farm transmission.

In conclusion, infected mink farms can be highly contaminated with SARS-CoV-2 RNA in airborne dust, on surfaces, and in various other environmental matrices. Dispersion of SARS-CoV-2 to outdoor air was limited, which implies a negligible environmental exposure risk for neighbouring residents.

Our occupational and environmental risk assessment supports earlier reported whole genome sequencing research showing mink-to-human transmission in farm workers but no direct zoonotic transmission events to nearby communities.

## Data Availability

The datasets generated during and/or analysed during the current study are available from the corresponding author on reasonable request.

## Author contribution

Conception and design: M.d.R., L.S., B.O.M., R.S., A.v.d.S., M.S., R.B., R-J.M., M.K., A.S., W.v.d.P.

Data collection and laboratory work: M.d.R., R.H.v.d.H., M.H., F.H., M.E., W.v.d.H., K.M., S.N., I.v.S., J.S., L.S.

Data analyses and interpretation: M.d.R. together with L.S. with additional input from R-J.M., M.K., A.S., W.v.d.P.

Preparation of manuscript: M.d.R. together with L.S. with additional input from R.H.v.d.H., M.H., F.H., M.E., W.v.d.H., K.M., S.N., B.O.M., I.v.S., R.S., A.v.d.S., M.S., J.S., R.B., R-J.M., M.K., A.S., W.v.d.P.

## Supplemental Information available online

Supplemental Methods (Parts A-C), Supplemental Tables (S1-S2), Supplemental Figures (S1-S3)

### Acknowledgements

We thank the farm owners for their cooperation. We would like to acknowledge Sophie van Oort, Wietske Dohmen, Angèle Timan and Monique Tersteeg-Zijderveld for their contribution to the laboratory analyses.

## Supplemental Information

### Supplemental Methods

#### Supplemental Methods A. Background information on investigated farms

The first SARS-CoV-2 infected mink farms were investigated thoroughly including sampling of animals, humans and the environment. These farms were all situated in the region of the Netherlands with the highest mink farm density, the eastern part of the province of Noord-Brabant (NB). On April 23^rd^, SARS-CoV-2 infection was established in the first farm, NB1, which has two separate locations, NB1A and NB1B, which are 115m apart. Two days later another farm was diagnosed positive, NB2, situated at 14 km distance from NB1. For NB1 as well as NB2, the source of the outbreak amongst minks was traced back to an SARS-CoV-2 infected farmer/worker at the specific farm. Results of whole-genome sequencing research indicated the virus strain at NB2 to be different from NB1, thus underlining the occurrence of two separate antropozoonotic transmission events^2,9^. Both NB1 and NB2 experienced increased mortality amongst the minks coinciding with respiratory signs starting in the first half of April but, as it was unprecedented, initially SARS-CoV-2 infection was not suspected. Serological findings suggested, based on a random subset of minks tested, many minks to already have seroconverted by the time of diagnosis, indicating the outbreak to be indeed circulating for a while. This was confirmed by sequencing research showing considerable viral genomic diversity at the time of diagnosis. Animal investigations performed from the moment of diagnosis onwards, indicated the number of infected minks to be decreasing over time thus substantiating that both NB1 and NB2 were in a later phase of the outbreak when detected. From 28^th^ of April onwards, environmental sampling started at and around NB1 and NB2. On May 6^th^, SARS-CoV-2 infection was detected at two more farms, NB3 and NB4. NB4 was, in contrast to the other farms, at the time of diagnosis in a more early phase of the outbreak based on disease history, serology and sequence diversity. Therefore, NB4 was included for environmental sampling from May 13^th^ onwards, while NB3 was not included. For a detailed description on the course of the outbreak in the farmed minks, clinical/pathological findings, and genomic epidemiology see Oreshkova et al.^2^, Molenaar et al.^10^, and Oude Munnink et al.^9^.

Minks kept at NB1, NB2 and NB4 were housed in wire netting cages placed in halls. Halls are naturally ventilated via both large openings in the roof as well as walls which are only (partially) closed in the winter months. Cages are arranged in long single rows, on one side separated by a narrow manure conveyer belt and on the other side by a feeding alley (see pictures in Supplemental Methods B). In the front of each cage is a sleep/nest box which contains bedding material. Solid boards are attached onto the sides of the cage to prevent direct contact between minks in bordering cages. Per cage, one adult mink is housed and if present kits are kept with their mothers. Whelping takes place once a year in the period end of April/beginning of May. The population before whelping consisted of: 8971 females and 24 males at NB1A, 2923 females and 1699 males at NB1B, 7500 females and 90 males at NB2, and 10300 females and 242 males at NB4.

##### Timeline of the outbreaks at the mink farms in 2020 and environmental sampling

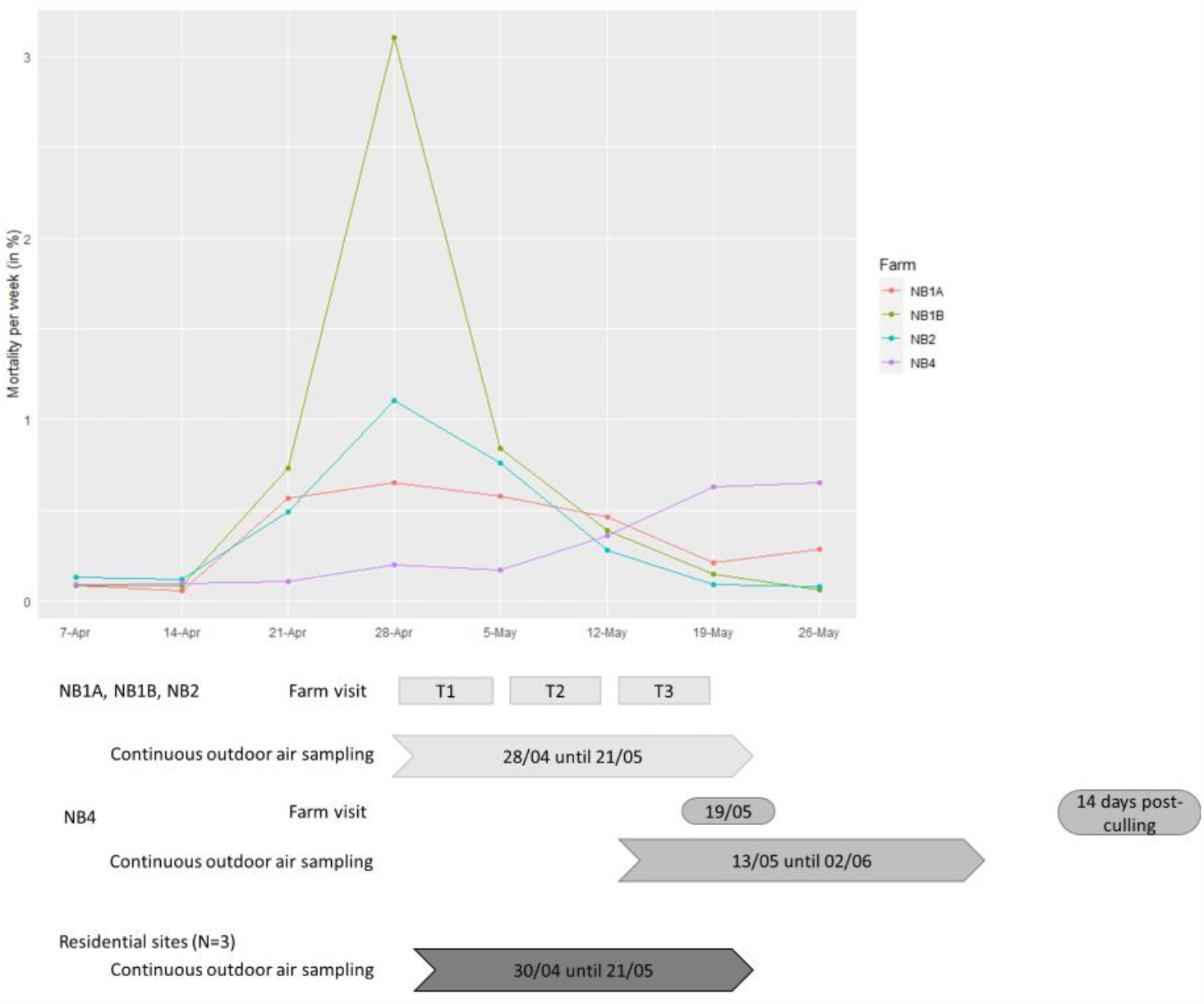

Note. T1: NB1A 28/04, NB2 30/04, NB1B 02/05 T2: NB1A 05/05, NB2 07/05, NB1B 09/05

T3: NB1A 12/05, NB2 14/05, NB1B 16/05

Mortality is expressed in percentage of naturally deceased minks per week. GD Animal Health is acknowledged for providing the data on mortality^10^.

### Supplemental Methods B. Additional information on sampling Technical details on air sampling

#### Outdoor air sampling

Multiple-day sampling of PM_10_ was performed by means of Harvard impactors (Air Diagnostics and Engineering Inc., Naples, ME, USA) which selectively sample PM_10_ by trapping larger particles on an impaction plate. Harvard impactors were connected to self-designed air pumps with critical orifices calibrated at a flow of 10.0 l/min (optimal flow rate for this sampling head type). To enable sampling of total suspended particles the impaction plate was removed from the Harvard impactor for a series of measurements at farm NB4. For inhalable dust sampling, GSP (Gesamtstaubprobenahme, total dust sampling) sampling heads were used connected to a Gilian GilAir 5 pump (Sensidyne, St. Petersburg, USA) calibrated at a flow of 3.5 l/min (optimal flow rate for this sampling head type). Sampling heads were attached side-by-side onto a pole at 1.50m height (average breathing height of humans; see Supplemental Methods for a picture of the measurement set-up).

#### Sampling inside farm and downwind/upwind

Six-hour stationary air sampling was performed and 8-hour personal air sampling by using Gilian GilAir 5 pumps with battery pack. Inhalable dust sampling was performed by means of GSP sampling heads at a flow of 3.5 l/min. PM_10_ sampling was performed by means of PEM (Personal Environmental Monitor) sampling heads (MSP Corporation, Minnesota, USA) at a flow of 4.0 l/min (optimal flow rate for this sampling head type) which selectively samples PM_10_ by trapping larger particles on an impaction plate. Stationary air sampling was performed by attachment of the sampling heads side-by-side onto a pole at 1.50m height (average breathing height). Personal air samples were collected by attachment of the sampling heads within the breathing zone of the fieldworker and the pumps clipped on a belt; see Supplemental Methods for pictures of the measurement set-up. Inside the farm, stationary air sampling was performed at three spots distributed within the farm which remained the same per farm. The spots of the six-hour air sampling outside the farm were based on the wind direction of the measurement day. Upwind sampling was performed at 50 meter distance from the farm, and downwind sampling at 10-20 meter and 100 meter distance.

### Pictures showing sampling of air and settling dust

Picture showing stationary air sampling (1) and sampling of settling dust in close proximity of minks (2).

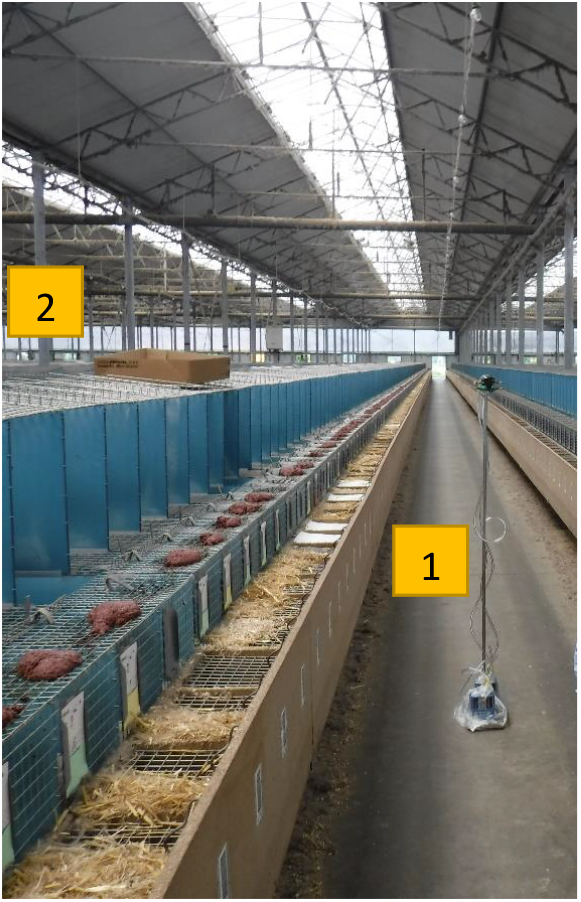

Close-up of EDC placed in cardboard box

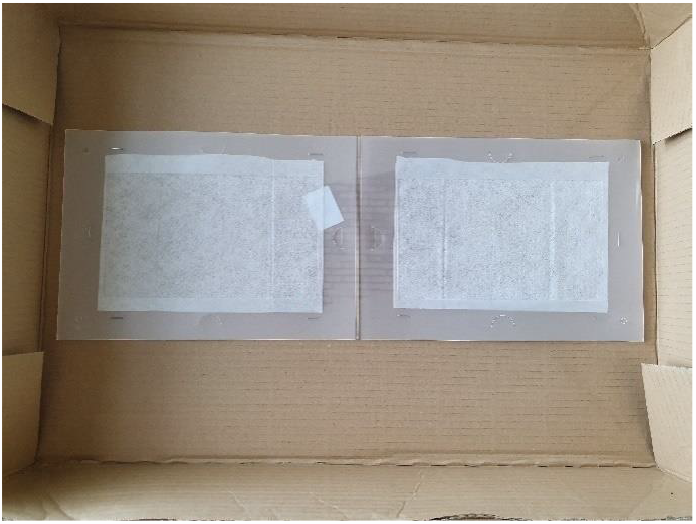

Pictures showing sampling of settling dust at further distances from minks; left by placement of EDC on hanging PVC plate (red circle), right by placement of EDC on stand in empty row

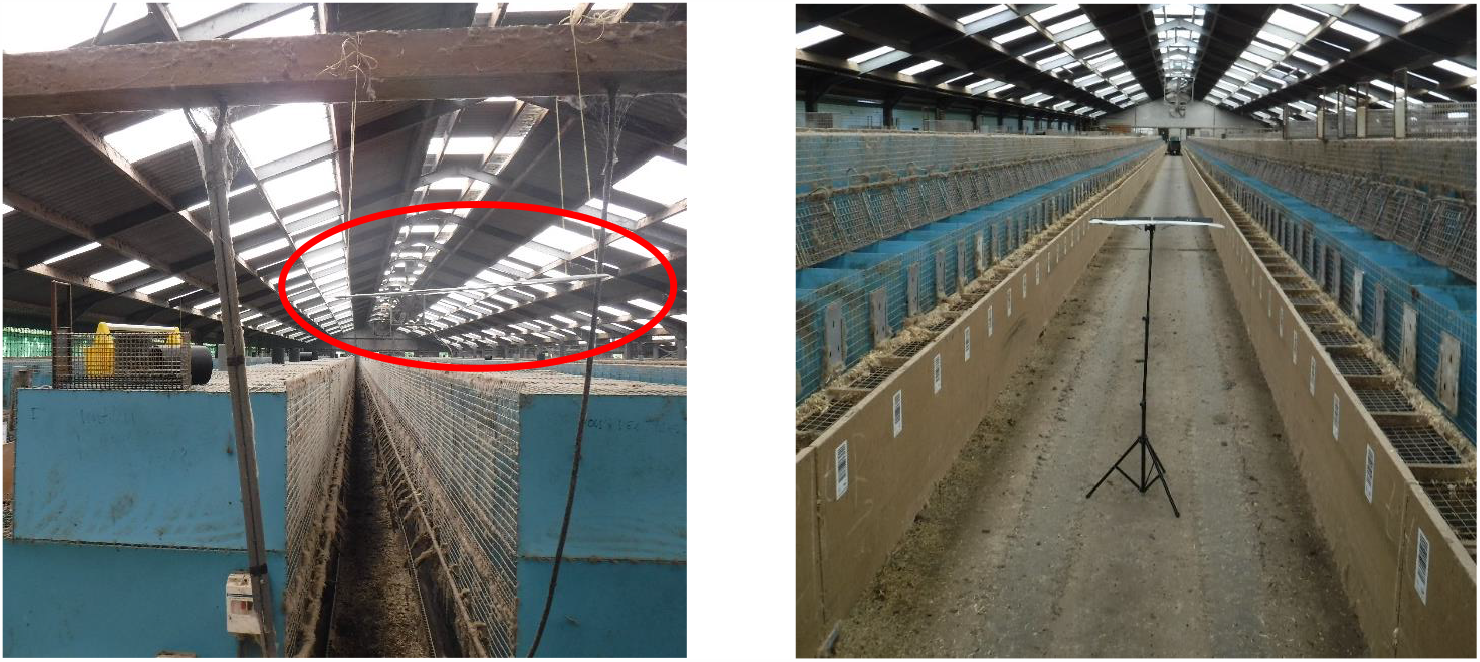

### Details on sampling of mink’s housing units

Picture of mink’s housing unit

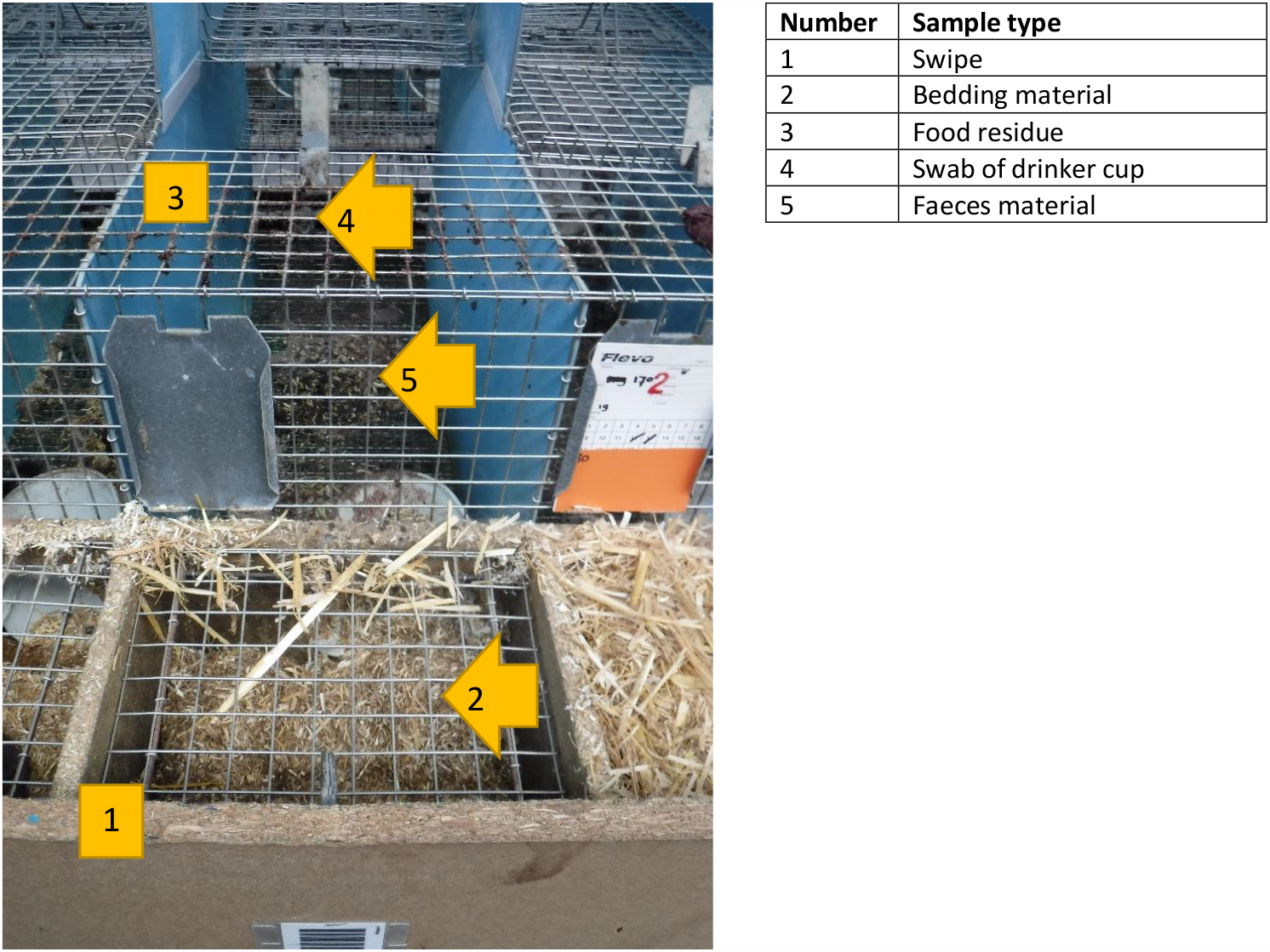

Picture showing typical way of feeding minks by placing suspension of raw meat on top of the cage

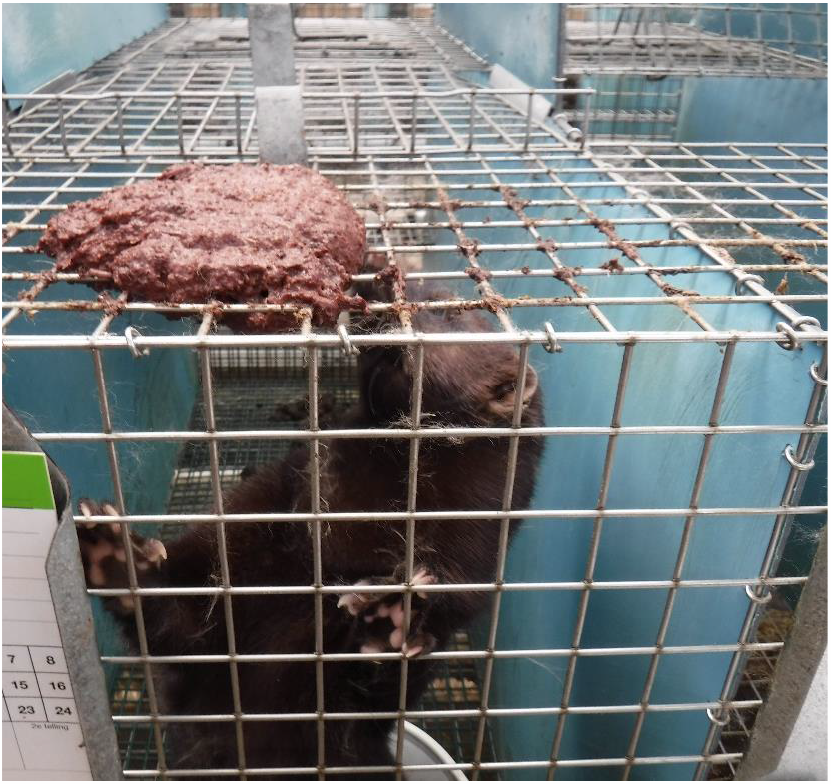

### 1. Collection of swipe

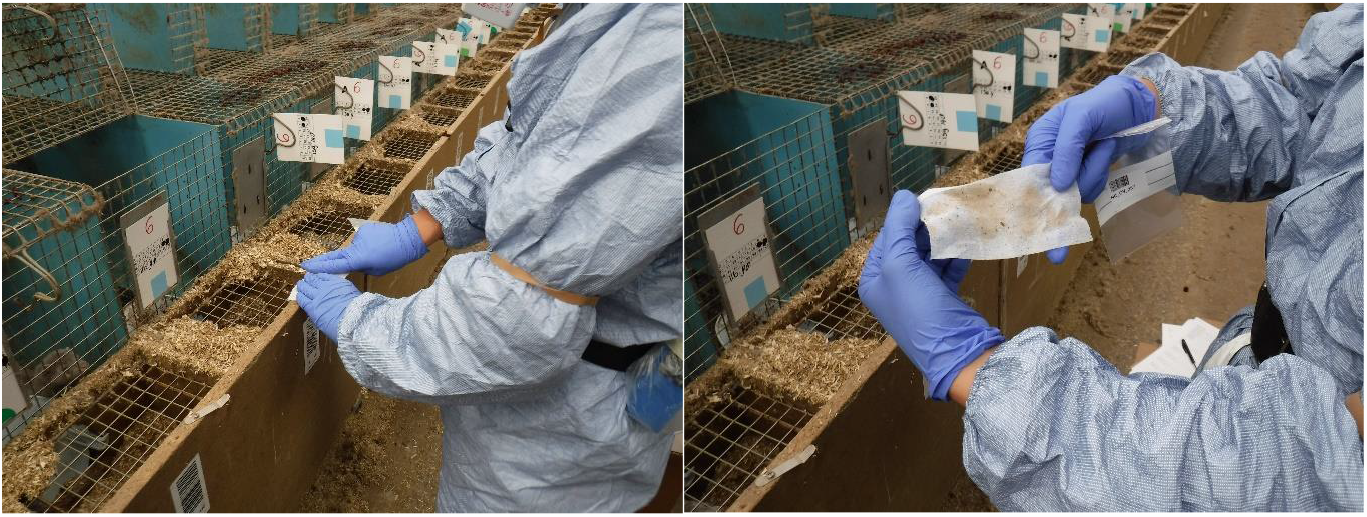

Use sterilized electrostatic cloth to collect material settled on hardboard rim. Swipe surface of 135mm length and 65mm width. Wear clean gloves before every sampling.

### 2. Collection of bedding material

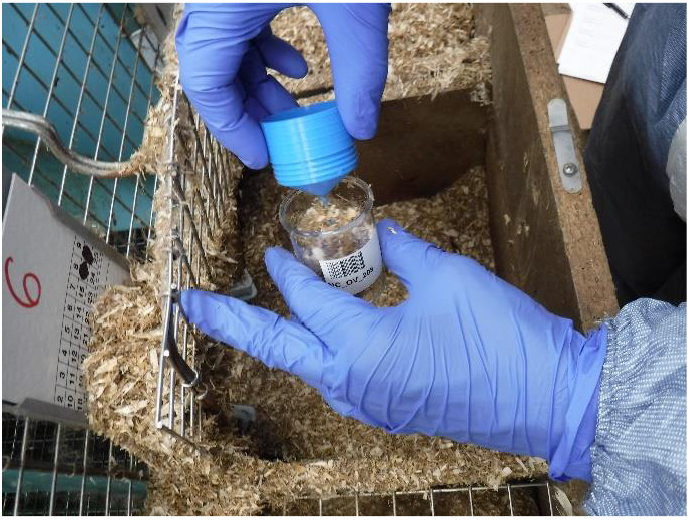

Collect bedding material from the night/nest box by scooping it in a container (sterilized, DNA/RNA-ase free).

### 3. Collection of food residue

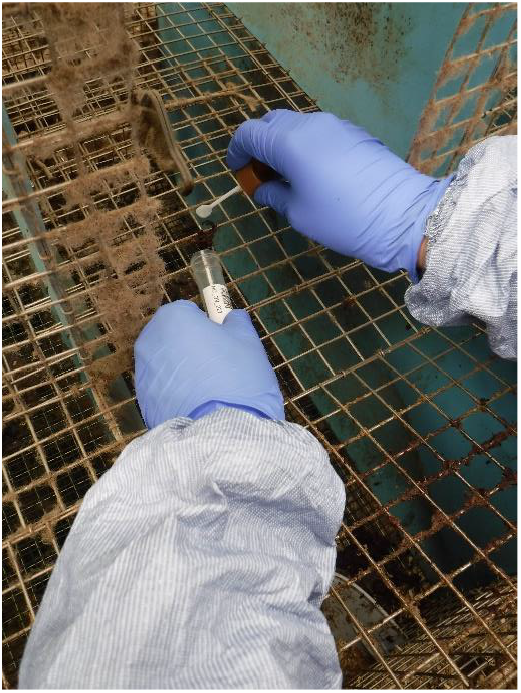

Scrape off the wire the residues of the food that are left and capture the residues in a container (sterilized, DNA/RNA-ase free)

### 4. Collection of swab of drinker cup

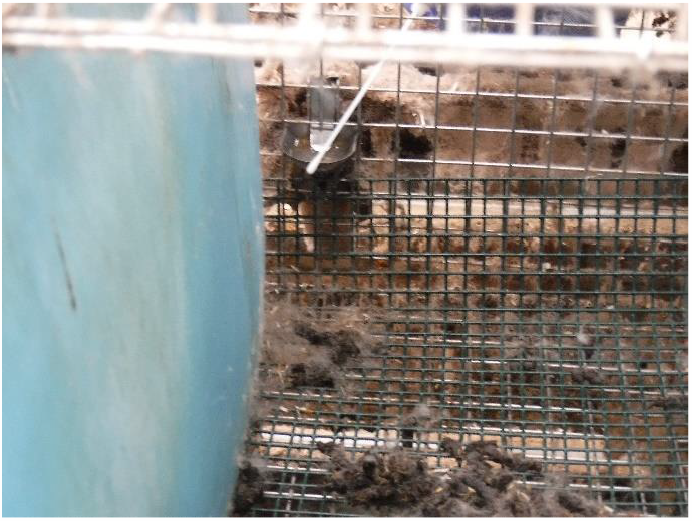

Swabbing of the drinker cup by rolling the swab (sterilized, DNA/RNA-ase free) on the rim.

### 5. Collection of faeces material

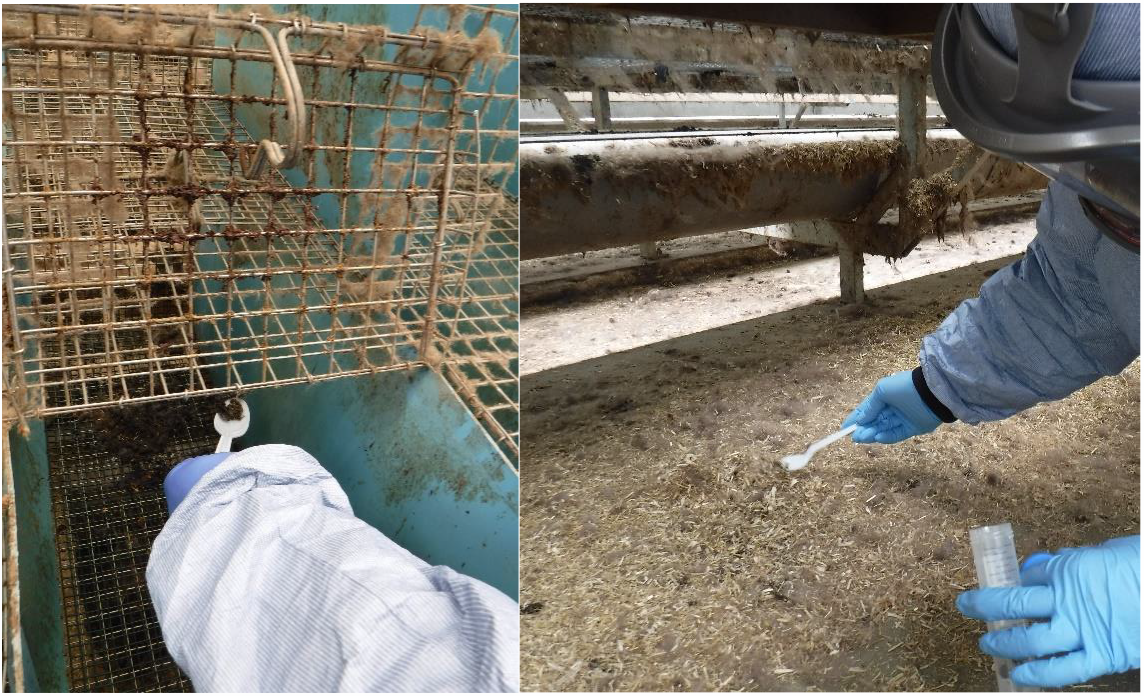

Collect faeces material from the cage when present or from the floor beneath the cage by scooping it in a container (sterilized, DNA/RNA-ase free).

### Supplemental Methods C. Details on laboratory analyses

#### Sample processing for storage at the end of fieldwork day

For the sampling heads used for air measurements, this involved disassembly to collect the exposed filter and transfer it to an enclosed tube (Greiner 15ml, DNA/RNA-ase free) but also taking a swab of the impaction plate used in PM_10_ sampling which enabled exploration of potential differences related to size fraction. For the EDCs, this involved collection of the exposed electrostatic cloth out of the holder and place it in an enclosed tube (Greiner 50ml, DNA/RNA-ase free). The other samples were stored directly in their collection container at -80°C.

#### Further processing for RNA extraction

Per sample type, an adequate processing procedure was performed.

The exposed Teflon air filters, stored at -80°C in 15 ml Greiner tubes with the dust-surface facing the inner of the tube were thawed in the safety-cabinet of the BSL-2 lab. Using a 5 ml sterile pipet 4 ml of PBS-Dulbeco’s was added. To submerge the complete filter of 37 mm in diameter in the fluid, the filter was pushed carefully, without damaging the Teflon surface, to the bottom of the tube using the tip of this 5 ml pipet. Tubes were placed on an end-over-end roller and after 90 minutes of rolling at room temperature the tubes were placed vertical for a few minutes allowing the fluid to drip to the bottom of the tube. Using a 2 ml pipet the fluid and large dust particles that sank to the bottom, were homogenised and 1 ml of the suspension was transferred to a clean Eppendorf tube. After vortexing the Eppendorf, 200 µl of suspension was mixed with 200 µl lysis buffer of the ID Gene™ Mag Fast Extraction Kit (ID-VET) and RNA extraction was performed on the KingFisher (ThermoFisher). The remaining suspensions were stored at -80°C for potential virus isolation.

EDC cloths stored at -80°C in 50 ml Falcon tubes with the swiped surface facing the inner of the tube were thawed in the safety-cabinet of the BSL-2 lab. Using a sterile pipet 10ml PBS-PEFABLOC buffer was added (PBS-PEFABLOC buffer: 1% v/v of stock solution in PBS-Dulbecco’s. Stock solution: 1 PEFABLOC tablet [Sigma Aldrich, Zwijndrecht, the Netherlands] dissolved in 1ml PBS-Dulbecco’s and stored in aliquots at -20°C) and rolled for 1 hour on an end-over-end roller. Two hundred µl of the suspension with dust particles was collected from each tube and mixed with lysis buffer in a similar manner as described above for Teflon air filters.

Approximately 0.5 ml bedding material was transferred to a 15ml Falcon tube to which 3ml PBS-PEFABLOC buffer was added. After vortexing, tubes were incubated one hour at room temperature and centrifugated 10 minutes at 2500xg. Two hundred µl of intermediary fluid was collected and mixed with 200 µl of lysis buffer for RNA extraction.

For food residue samples as faecal material, approximately 0.5 ml was transferred to a 15ml Falcon tube. To each tube 2 ml PBS-PEFABLOC buffer was added and tubes were vortexed vigorously until the material was properly suspended. After incubation of one hour at room temperature, tubes were centrifuged for 10 minutes at 2500xg rpm and 200 µl of the supernatant was collected and mixed with 200 µl of lysis buffer for RNA extraction.

Sampled swabs (both swabs taken from drinker cups as well as impaction plates) were transferred to a 15ml Falcon tube with 2 ml of medium used for maintaining Vero-E6 cells (Minimum Essential Medium with 5% v/v foetal bovine serum (Gibco, Thermo Fisher Scientific, Bleiswijk, The Netherlands), 1% v/v Antibiotic-Antimycotic mixture (anti-anti, Gibc0), 1% L-Glutamine (Gibco), 1% nonessential amino acids (Gibco). Tubes were vortexed and incubated for one hour at room temperature. After centrifugation for 10 minutes at 2500 g, 200 µl of the supernatant was collected and mixed with 200 µl of lysis buffer for RNA extraction.

#### PCR

Subsequently, samples were tested for SARS-CoV-2 using the accredited E gene PCR as described by Corman et al.^26^ using the TaqMan Fast virus 1-Step Master Mix (Applied Biosystems), with minor modification in the reverse transcription conditions (52°C for 10 min). A calibration curve was established based on serial dilutions of inactivated cell culture produced SARS-CoV-2 with known titer (SARS-CoV-2/human/NL/Lelystad/2020). Samples of the same type were tested in one or maximally two runs. Negative controls (no template) and positive controls were included in each PCR run. To each sample, an internal control (intype IC-RNA, Qiagen) was added to check for inhibition of reverse transcription and amplification. Samples with Ct values below the threshold Ct, were defined positive. The threshold Ct was set at 36 based on optimization of sensitivity and specificity analysed by means of the ROC curve using the function spEqualSe from R package OptimalCutpoints.

#### Attempted virus isolation from air filters

After establishing the presence of SARS-CoV-2 RNA by E gene qPCR in air samples, samples with a Ct value below 32 were subjected to virus isolation using Vero-E6 cells. After thawing, 400 hundred µl of suspension was used to infect monolayers containing of 1×10^6^ Vero-E6 cells (ATCC) grown in T25 cm^2^ flask. Cells were cultured in MEM supplemented with 5% FCS, 1% antibiotic/antimycotic, 1%, glutamine and 1% non-essential amino acids at 37°C and 5% CO2. Monolayers were inspected regular with a microscope to identify the typical cytopathogenic effect induced by SARS-CoV-2. After five days of growth 200 µl of the medium was analysed by E-gene qPCR to detect replication of SARS-CoV-2.

##### Schematic overview of PCR analysed proportion per sample type

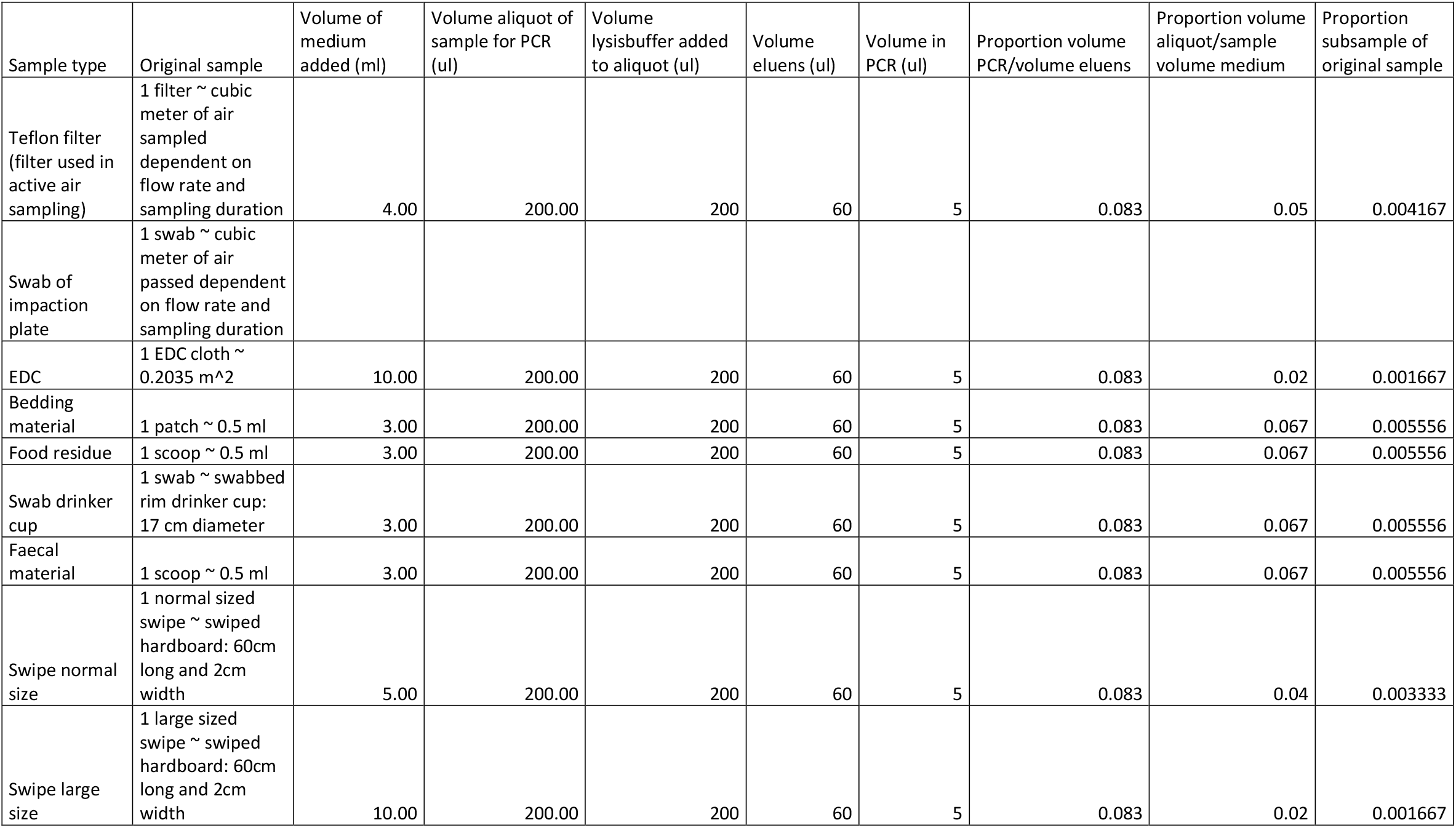

## Supplemental Tables

**Supplemental Table S1.** Results of multivariable modelling on SARS-CoV-2 RNA detection in settled dust sampled by means of EDCs.

|  | Virus load (log copies) |  |  |
| --- | --- | --- | --- |
|  | EDC |  |  |
| Variable | Ratio | 95% CI LB | 95% CI UB |
| Farm NB1A (indicator) | . | . | . |
| Farm NB1B | 0.41 | 0.16 | 1.00 |
| Farm NB2 | 18.00* | 6.70 | 50.00 |
| Timepoint 1 (indicator) | . | . | . |
| Timepoint 2 | 0.28* | 0.12 | 0.67 |
| Timepoint 3 | 0.05* | 0.02 | 0.12 |
| Positioned in close proximity to minks | 3.00* | 1.20 | 7.50 |

Note. Ratio = estimate of association to the power 10 to represent ratio in viral load 95% CI LB = lower bound of the 95% confidence interval 95% CI UB = upper bound of the 95% confidence interval Marked \* then P-value < 0.05

**Supplemental Table S2.** Comparisons of SARS-CoV-2 RNA detection in 14 housing units at NB4 sampled pre-culling and post-culling.

| Sample type | % positive samples pre-culling | % positive samples post-culling |
| --- | --- | --- |
| Swipe | 100% | 14% |
| Bedding material | Mix of layers: 100% | Bottom layer: 85%<br>Top layer: 57% |
| Faeces material | 71% | 21% |
| Swab drinker cup | 50% | 0% |
| Food residue | 14% | 0% |

## Supplemental Figures

**Supplemental Figure S1.**
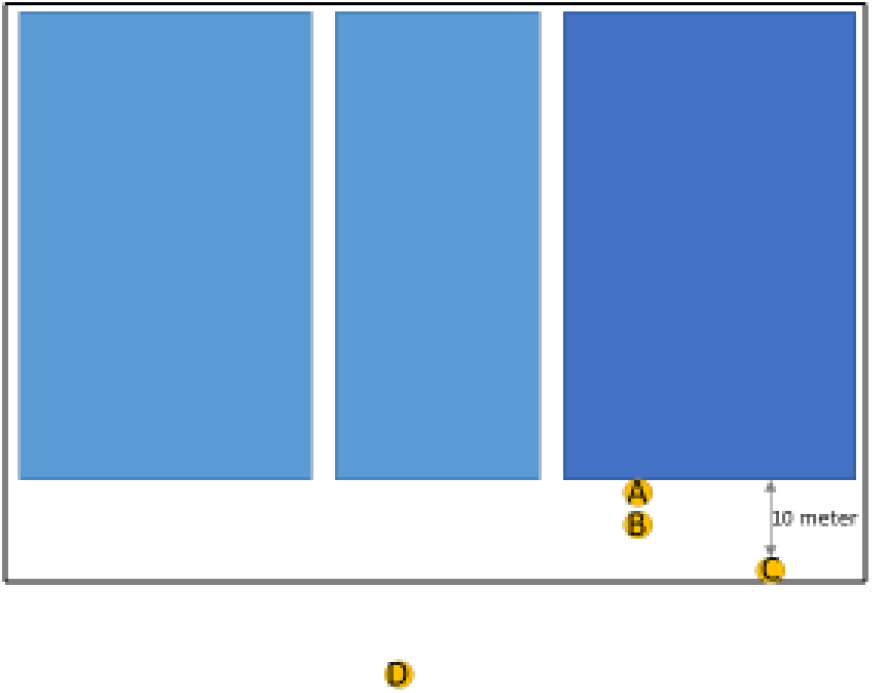
Map showing the sampling locations at NB4. The blue blocks are mink stables.

**Supplemental figure S2.**
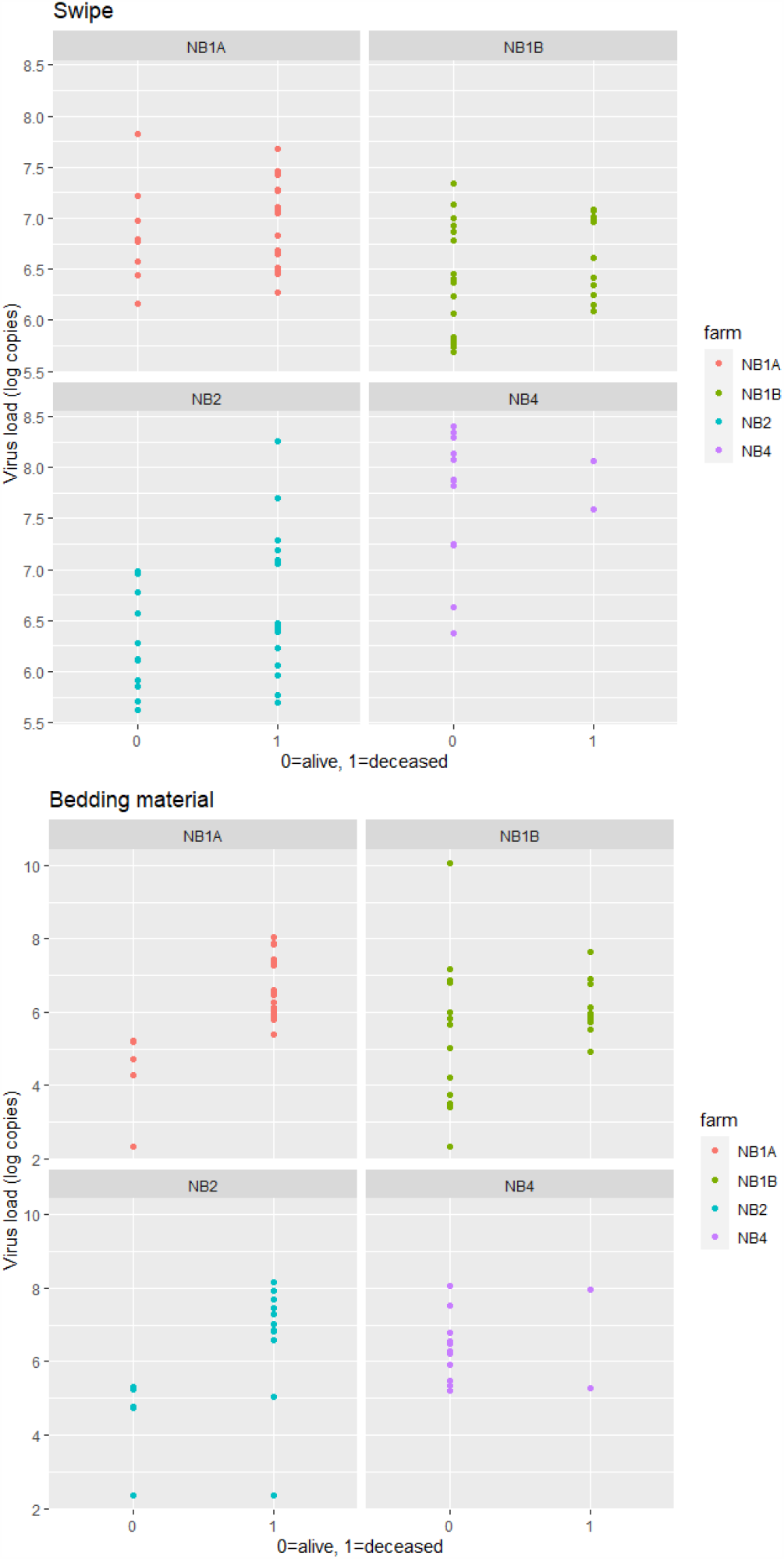
Overview of measured viral load in swipes and bedding material samples per farm in housing units belonging to recently deceased minks versus minks still alive.

**Supplemental Figure S3.**
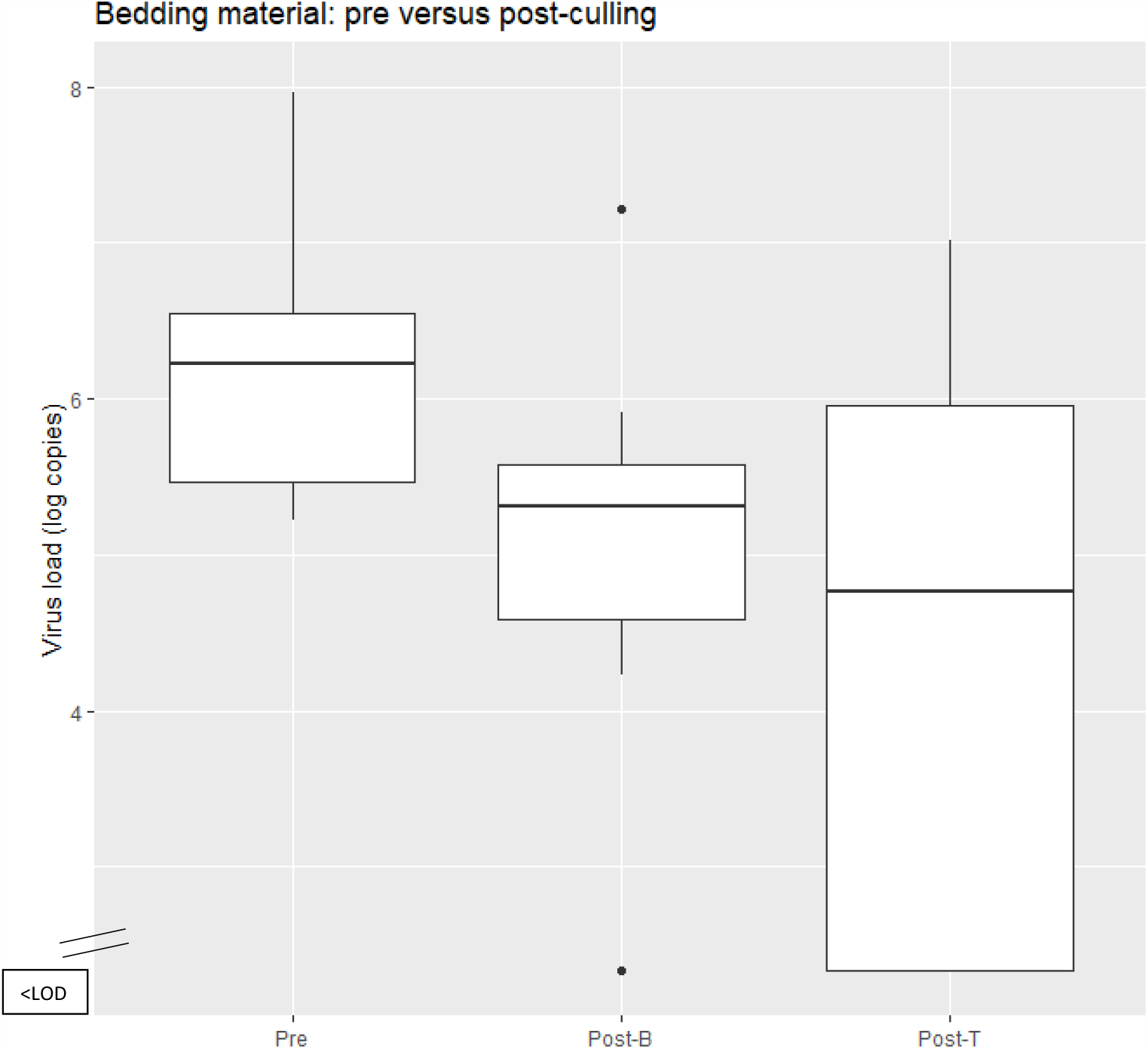
Overview of viral loads in bedding material collected pre-culling versus post-culling (divided in bottom layer sampling and top layer sampling) Note. Pre = bedding material samples collected pre-culling Post-B = bedding material samples collected post-culling of bottom layer Post-T – bedding material samples collected post-culling of top layer Percentage of samples <LOD, pre-culling 0%, post-culling bottom layer 15%, post-culling top layer 43%

